# Novel Approach for Monte Carlo Simulation of the new COVID-19 Spread Dynamics

**DOI:** 10.1101/2020.12.03.20243220

**Authors:** Stavros Maltezos, Angelika Georgakopoulou

## Abstract

A Monte Carlo simulation in a novel approach is used for studying the problem of the outbreak and spread dynamics of the new COVID-19 pandemic in this work. In particular, our goal was to generate epidemiological data based on natural mechanism of transmission of this disease assuming random interactions of a large-finite number of individuals in very short distance ranges. In the simulation we also take into account the stochastic character of the individuals in a finite population and given densities of people. On the other hand, we include in the simulation the appropriate statistical distributions for the parameters characterizing this disease. An important outcome of our work, besides the generated epidemic curves, is the methodology of determining of the effective reproductive number during the main part of the new daily cases of the epidemic. Since this quantity constitutes a fundamental parameter of the SIR-based epidemic models, we also studied how it is affected by small variations of the incubation time and the crucial distance distributions, and furthermore, by the degree of quarantine measures. In addition, we compare our qualitative results with those of selected real epidemiological data.

## 1 Introduction

Facing the problem of the spread of the new COVID-19 disease, the standard procedure is to analyse or to fit the epidemiological data with the appropriate mathematical models. This methodology is very useful, not only to determine the particular parameters of the disease, but also to forecast the evolution of its spread by using some typical figures of merit such as the doubling time, the peaking time and the basic and affective reproductive numbers [1, 2, 3, 4]. Another approach is to work in the reverse problem, that is, to generate epidemiological data consistent with the disease under study. By data we mean the fundamental ones, that is, the reported - confirmed “daily new cases” (DNC), expressing the daily rate of infected individuals [5]. The advantage of this methodology, implemented by a Monte Carlo (MC) simulation, is that it allows the use of the stochastic character of the parameters used in the mathematics of the epidemics (the known classical SIR model and its extensions), as well as including other extrinsic factors, like quarantine, physical distances and other specific social measures. In this methodology we are studying the set of people in a city or even in a country as a complicate system in which there is no sense in concentrating on to the behaviour or habits of individuals or groups of them in detail. Besides, this is surely infeasible. Two factors only must be taken to account, the first is the radius of movement or transport and the second the crucial physical distance for transmitting the disease. The choice of the appropriate range of values, because we are facing random variables with the associated distributions, is one of the main tools for the manipulation of the generated data. To be consistent with the new COVID-19 disease we have used medical data which can be found in available literature. However, the spread of the disease differs widely among the countries and thus we are forced to focus on the particular ones with a similar epidemic “picture”. Another very important and useful outcome of this simulation is the so called “effective reproductive number”, *R*_e_, which is a function of time. From this we can determine also the fundamental parameter used in SIR-based models, the “basic reproductive number”, *R*_*o*_. Both quantities present very large uncertainties when they are calculated from real epidemiological raw data (the DNC), even in the case of using parametrization mathematical models [6, 7, 8, 9, 10]. The reason for this is, firstly, the large fluctuations of the data and secondly the mathematical procedure for determining *R*_e_ based on variations day-by-day, as we describe in the following Sections. Studying the impact of the main parameters of the disease to *R*_e_ or its time trend is also a basic subject of our work and we suspect that the results can be useful for understanding and facing this serious worldwide pandemic.

## 2 Simulation methodology

The Monte Carlo (MC) simulation we have developed is based on the idea of simulating the natural mechanism and process which occurred during the spread of the disease [11]. In accordance with our claim that we must examine and study the community as a complicated system, we clarify the basic conditions by assuming the following: a) the transmission of the virus occurs only if the physical distance becomes small in the sense of it is in a crucial range where the probability of transmission tends to be equal to one asymptotically, b) the step of movement of a group of people during the day is a free variable of the simulation because it depends on personal or social factors and c) the specific parameters of the particular disease of new COVID-19 which are mainly, the incubation time and the recovering time, are combined by using a convolution [12, 13, 14, 15, 16, 17]. The three aforementioned conditions constitute the core of the simulation and thus their mathematical description is essential. In particular, due to their stochastic character we must determine their probability distribution and the appropriately chosen associated parameters, mainly the mean and standard deviation values. In addition, the population and the corresponding people density which are both related to what is called “size” in the mathematical theory of epidemics, must be specified at the beginning of the simulation. These conditions can be adapted to urban or country cases in their implementation in real conditions. Below we describe these conditions in more details, where the first three constitute random processes while the fourth one constitutes a set of given constant parameters [18, 19, 20].

- Daily step of movement (random process): the movement of the dots, because they represent real individuals, must present realistic characteristics. This step represents either a walking distance or longer distance (by using vehicles). However, because the movements are performed relative to the previous location, the cumulative distance should be much greater than one step. This parameter has been chosen to be a fraction of the length corresponding to the dimension of a grid shell according to the selected people density and follows a Gaussian distribution. In a typical - baseline run we set a mean step equal to 1/4 of the grid dimensions with a std equal to 1/12 of the grid dimensions. In Fig. 1 an indicative sequence of random movement by using discrete steps of two individuals during three successive days is presented.
- Physical distance (random process): the physical distance is a pivotal issue, in the sense that it is the fundamental parameter causing infection among two individuals in a short distance, if one of them is already infected. In order to introduce this condition in a realistic way, we must answer two fundamental questions, a) what is the appropriate relationship between the infection probability and the physical distance, and b) what is the statistical distribution and its parameters describing the variance of the probability related to this medical phenomenon. Even if an appropriate distribution is specified, the phenomenon in the real world is very complex: we can imagine the pair of individuals; one of them having a high or low virus load, not only keeping a shorter or longer physical distance, but also wearing a mask or not, or wearing it correctly or incorrectly, facing one another or not and other circumstances playing their particular role in the probability of infection. Moreover, the environmental conditions (temperature, relative humidity, air ventilation etc.) might contribute to the transmission probability. For this reason, the crucial parameters in the mathematical approach must be considered to be the more effective ones incorporating all the cases described above in a good approximation. The success of this approximation can be verified only in comparison to real epidemiological data analysis. The probability distribution that we assumed is the exponential one, whose the probability density function (PDF) is

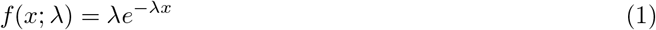

where *x* is a random variable (assumed *x* > 0) and *λ* is the “rate parameter”. The mean value is equal to 1*/λ* which is used in the trial runs of our simulation. As an alternative probability distribution that we could assumed is a sigmoid-shape function. In particular, we created a modified sigmoid function which can take values from maximum equal to 1 (for *d* = 0) to minimum equal to 0 (for *d* = ∞). This presents a saturation at very low values of the distance close to zero. If the corresponding probability distribution of infection approximate better or not is very hard to prove. However, this function has been tested by using an algorithm in the simulation code. This modified sigmoid function, assuming *d* ≥ 0 as the physical distance is the following

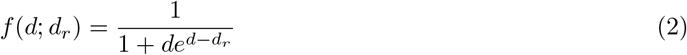

where *d*_*r*_ is a reference parameter specifying in which distance we want to achieve a particular probability of infection. For instance, setting *d*_*r*_ = 0.1931 the probability of infection at a distance of 0.5 m is equal to 0.5. For the typical physical distance of 2 m the probability decreases to 0.0528 (or about only 5.3%). In the present study of the MC simulation we have used the exponential distribution with mean value 8 m.
- Incubation and recovering time (random process): these parameters are very crucial for a realistic simulation. We are basically forced to use mathematical principles because of their stochastic character, due to the fact that both are related to medical processes inside a human body. It is known that the problems of waiting times are faced using the exponential distribution while in a more advance approach a Gamma distribution is assumed, which is belonging to the same generic distribution family. It is di-parametric including the positive parameters, shape *α* and the rate *β*. The corresponding PDF is the following.

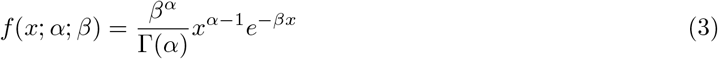

where *x* is a random variable, *x* ∈ (0, ∞). The mean value is *µ* = *α/β* and the variance *σ* ^2^ = *α/β*^2^. In our MC simulation, measuring the random variable of discrete time in days, we have chosen *a* = 6 (dimensionless) and *β* = 2*/*3 days^−1^. Therefore, the mean value is 9 days incorporating also the “latent period” and the rms (square root of variance) is 4.9 days. These two parameters lead to a distribution which show characteristics consistent with most medical observations published in the literature. In Fig. 2 we present an abstract design of the transmission mechanism between two individuals as a function of time, where one of them is infected and the other is considered as susceptible. We have to face a random process and thus we investigated the most realistic assumption for the required distributions.
- Population, Area and Density (given constants): in the simulation, we chose the total number of dots in a finite area. Therefore, the density is specified at the same time. The population density can be considered in large and very dense cities (e.g. like Paris) with around 20000 people per km^2^ or for typical small size cities with a density around 2000-4000 people per km^2^. In our trial runs we used the density of 2000 people per km^2^. As an initial condition we have chosen 10 infected individuals, constituting only 0.5% of the population.

**Figure 1:**
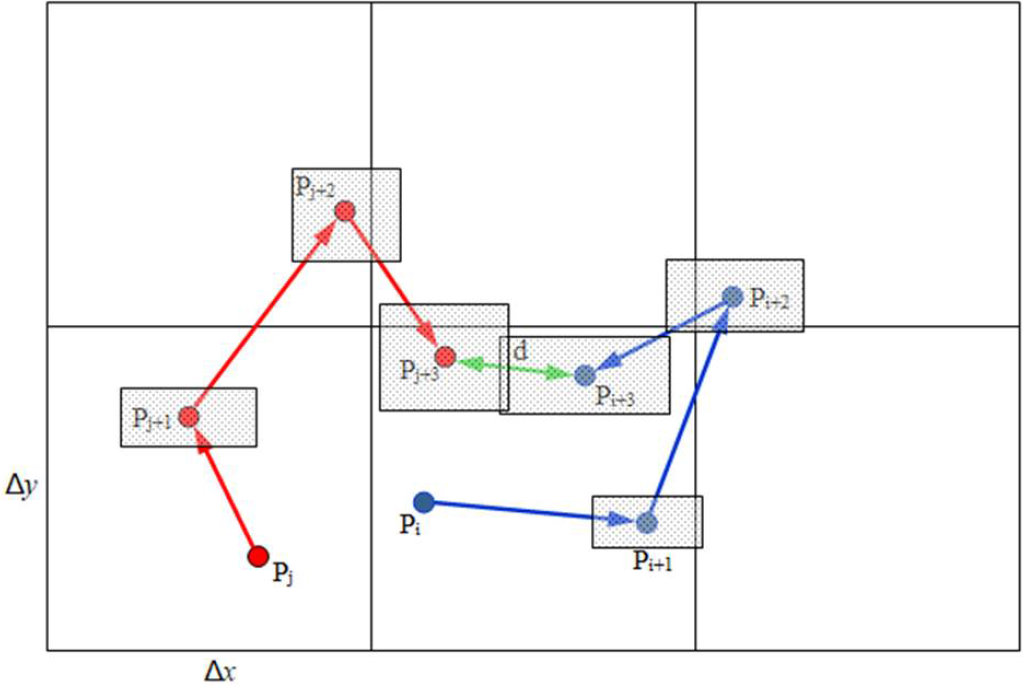
A random movement of two individuals *P*_i_ and *P*_j_ along a 2-D grid during three successive days. Their distance at the 3rd day is equal to *d*. The shade regions around any new location represent the uncertainty of the radial translations following Gaussian distribution in x and y coordinates.

**Figure 2:**
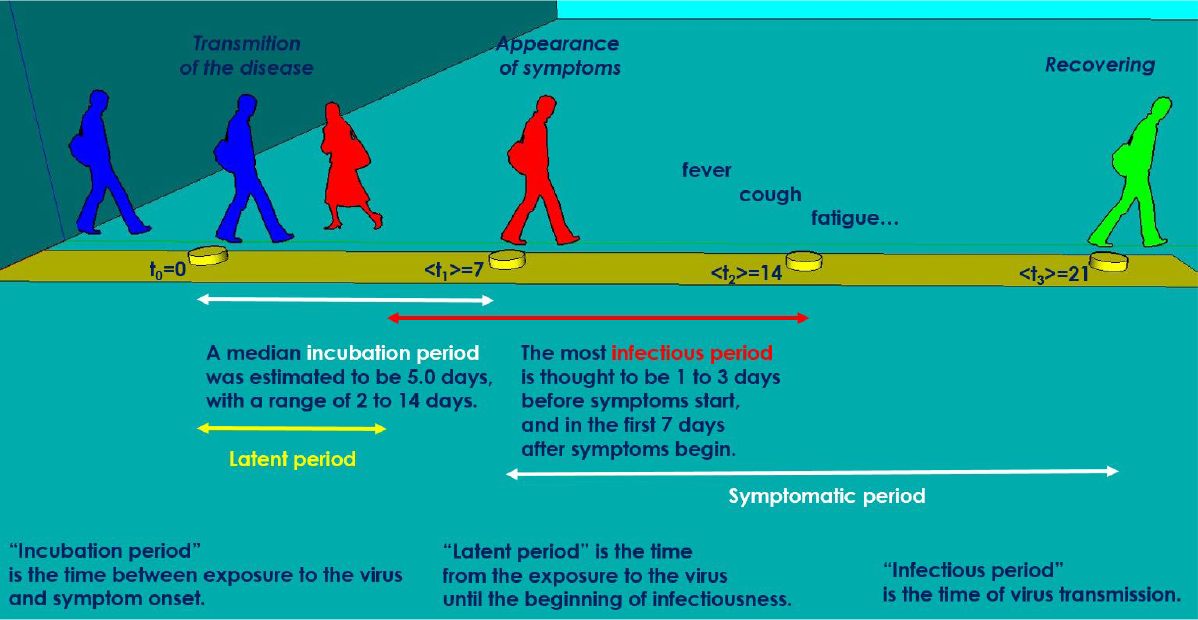
An abstract design showing the various time periods of the virus transmission mechanism from one infected individual to another. The time moments concern the mean values of the relevant random processes.

## 3 Determination of the effective reproductive number

The effective reproductive number, *R*_e_ is a fundamental quantity depending on the virus spread rate and the average number of contacts of individuals as a function of time. In particular, it expresses an average of how many secondary individuals one infected individual would transmit the virus to, during the mean infectious period. Theoretically, it can be determined in the frame of the SIR model as follows [21, 22, 23, 24, 25]:

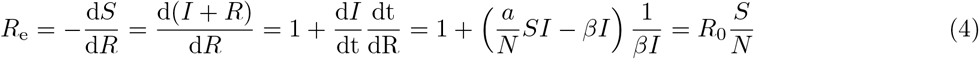

Because the condition for creating an epidemic is *R*_e_ *>* 1, the corresponding condition should be *S/N >* 1*/R*_0_. Also, at *t* = 0 should be *R*_e_(0) ≡ *R*_0_, at the peaking time *t* = *t*_p_ should be *R*_e_(*t*_p_) = 1 and at *t* = ∞ takes the value 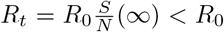. By using the expressions of Eq. 4 and using the third equation of SIR model 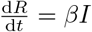, we can determine *R*_e_ at any time *t* based only the function *I*, as below

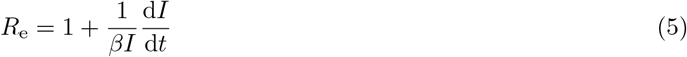

The generated data from the MC simulation are not only the DNC corresponding to the theoretical function of *t, I*, but also that corresponding to the theoretical functions *S* and *R*. The determination of *R*_e_ requires only the knowledge of *I* and as well as the parameter *β*. This parameter can be considered to be constant because it constitutes an intrinsic characteristic of the disease under study and can be available in the literature. Nevertheless, having the raw epidemiological data of the epidemic spread in a country, the value of *β* could be fitted or adjusted in order to be consistent with that of the daily recovered cases of individuals, let us call DRC. Even though these reported data are reliable, contain the relevant delay, in average 14 days, with respect to DNC ones. For this reason we prefer to adhere to the theory described by SIR-based models.

Below, we present the methodology to estimate the uncertainty of *R*_e_. According to Eq.5 the sources of the uncertainty are two, a) the uncertainty of *I* due to its statistical fluctuations and b) the deviation of constant parameter, *β*, from a standard bibliographic value. However, the latter, is of secondary importance because, first a hypothetical deviation is expected to be relatively small and secondly because an appropriate “fine tuning” can be performed. Therefore, in our calculations below we assume a fixed value for *β*.

In order to calculate the error’s transmission from *I* to *R*_e_ we can assume a short time period around *t* = 0, Δ*t*, where *R*_e_ can be considered constant. This assumption is realistic, and we can also say necessary, because the recovering rate in this disease is finite and the variations can be only be calculated within this time period. Being *R*_e_ constant, we can write

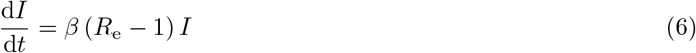

We integrate from an arbitrary time *t*_*i−*1_ to *t*_*i*_.

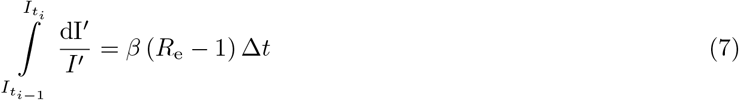

Leading to the expression

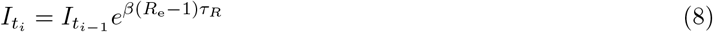

In the general case we can set *c* ≡ *βτ*_*R*_, where *c* differs slightly from one in practice. As we can observe, according to this assumption of constant *R*_e_ in finite time slots, its value depends on the logarithm of the ratio of the two successive values of *I* at the beginning and the the end of this time slot. This expression can also be very useful when a surveillance of the epidemic must be implemented for making a reliable prediction of it, mainly during its mitigation stage. At this point, based on this equation, Eq. 8, we proceed to solve the algebraic equation for *R*_e_ obtaining

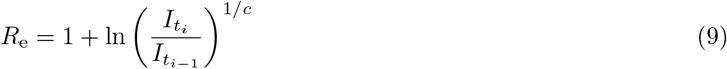

The values of *I* follow the Poisson distribution, presenting the associated uncertainty 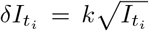 and 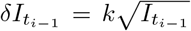 respectively, where *k* is a positive constant which is used for the cases of observing larger fluctuations in the raw data of *I* taking a value greater than one. Therefore, the uncertainty of *R*_e_ should be

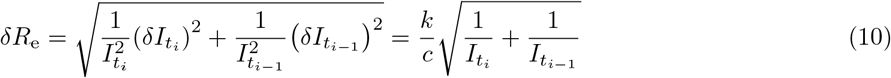

While the corresponding relative one is

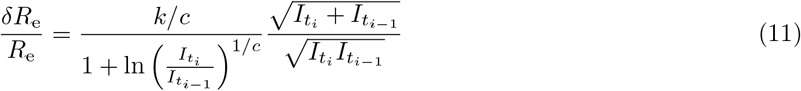

In the segment around the peak of the DNC curve, we theoretically expect *R*_e_ = 1. Indeed, from Eq.11 we obtain this value because of the maximum of the curve (having an extremum), where 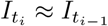. In this region, the relative uncertainty takes its minimum value and becomes

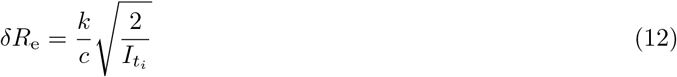

The basic reproductive number, *R*_0_, can be calculated by the above equations by using the values in the first time slot (typical is of the order of 13 days), that is, *I*_0_ and *I*_*t*_1. Definitely, *R*_0_ is the maximum value of *R*_e_, as it can be easily proved from Eq. 4 where at *t* = 0 the fraction *S/N* takes its maximum value and then is decreasing due to monotonically decreasing of *S*.

Implementing the above methodology in the simulation program we use two processes for improving the accuracy of *R*_e_ determination. The first concerns a digital filtering, reducing the statistical fluctuations, and the second is a moving average algorithm, both realized by functions of Matlab [26].

## 4 Implementation and results

### 4.1 Results obtained by the simulation

During a typical run of the MC simulation, the *N* dots within the assumed area are categorized by using colors: a) the black for the susceptible individuals, b) the red for the infected individuals and c) the green for the recovered individuals. The dots change their location day-by-day representing the random movement of the individuals in the real world. In Fig. 3 we present a diagram with the daily new data (DNC) generated by the MC simulation. In Fig. 4 the corresponding effective reproductive number calculated by the MC simulation as a function of time (red circles and spline solid line) is shown. We also used a smoothed curve (green solid line) by calculating the “moving average” every 13 days, which has been chosen as the optimum time interval. Focusing on the peaking time in both plots, we observe that the value *R*_e_ = 1 corresponds to the peaking time, as one might expect according to the SIR-based models. Moreover, apart from the gradually decreasing trend, we observe some slow variations, before and after the value corresponding to the peak, a phenomenon which is hard to be explained in a simple way. These variations are visible also in the obtained *R*_e_ from real epidemiological data (see 4.2). The maximum value is around 1.80 and the minimum, relatively adequate accurate value, is at the level of 0.2 around day 40.

**Figure 3:**
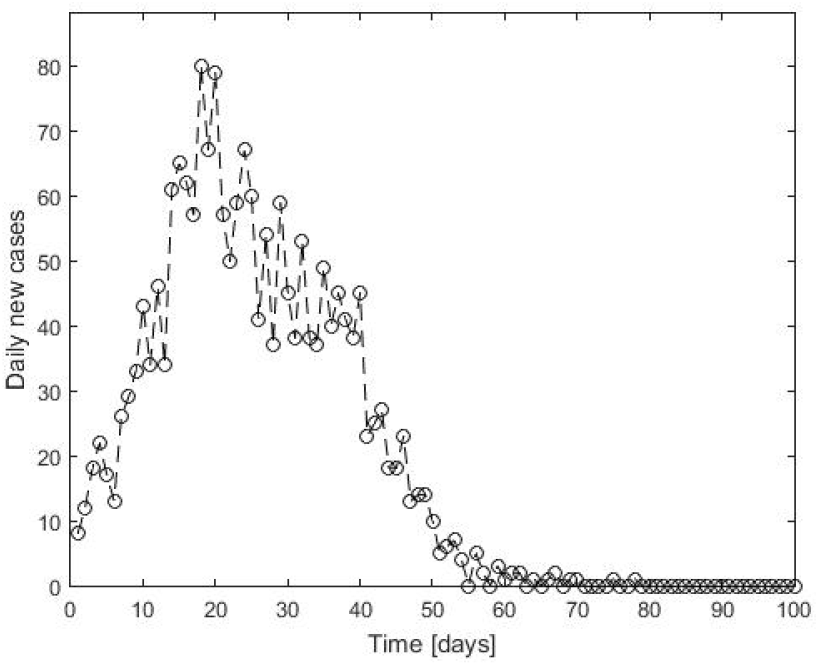
The generated epidemiological data (DNC) by the MC simulation, counted day-by-day during the run and cumulated along the whole area of movement.

**Figure 4:**
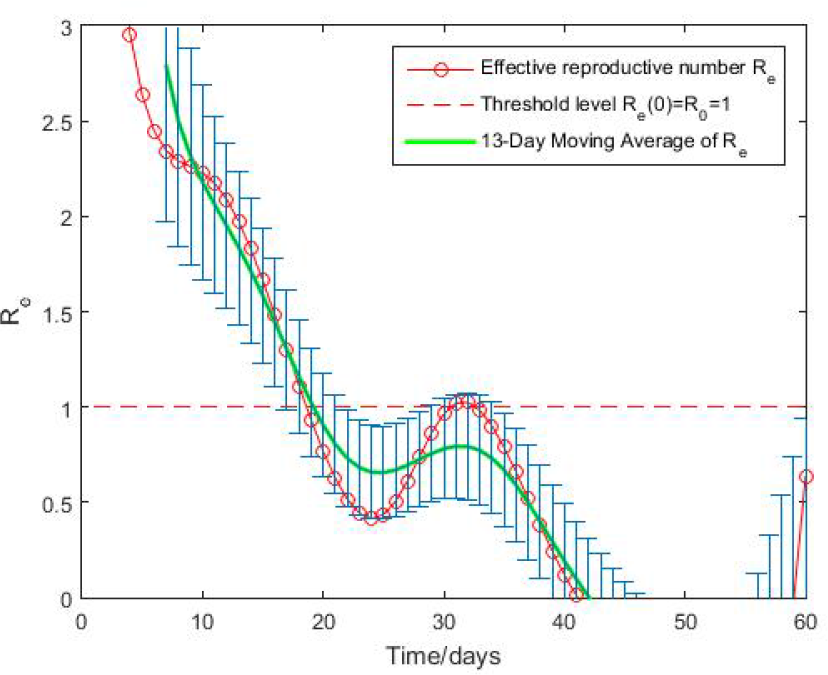
The effective reproductive number from the MC simulation as a function of time. The error bars correspond to the moving average values.

### 4.2 Qualitative discussions and comparisons

By the MC simulation we have studied the effect of the degree of quarantine, by means of the positive effect on the spread dynamic of the epidemic. We have set three degrees of quarantine by reducing the movement range of the individuals and the rms deviation in steps of 0.25, from zero (no quarantine) to 0.75. Because of the fluctuations caused by the stochastic process of the MC simulation we were running the MC simulation 10 times and calculating the mean and rms value for each particular quantity. In Fig. 5, 6, 7 and 8 these results are plotted with the associated fitted curves. In Table 1 the obtained numerical results are summarized.

**Table 1:**
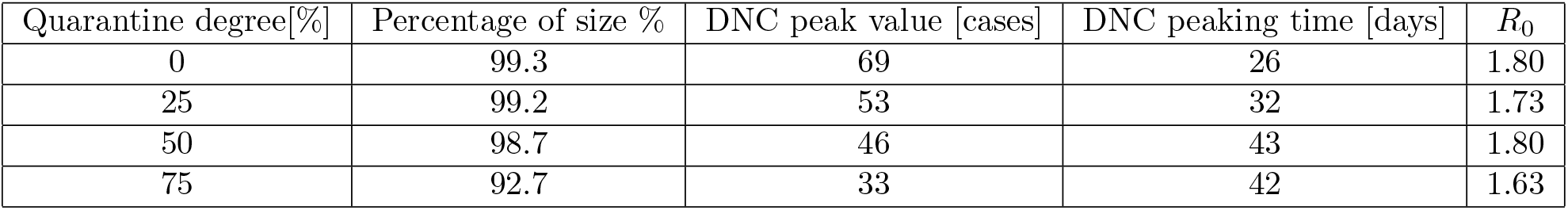
Summary of the MC simulation results related to the effect of the quarantine degree, with the same density and the mean daily step and the STD of daily step was that of the baseline run, that is, 7.90 m and 2.64 m respectively. The density was set 2000 p*/*km^2^.

**Figure 5:**
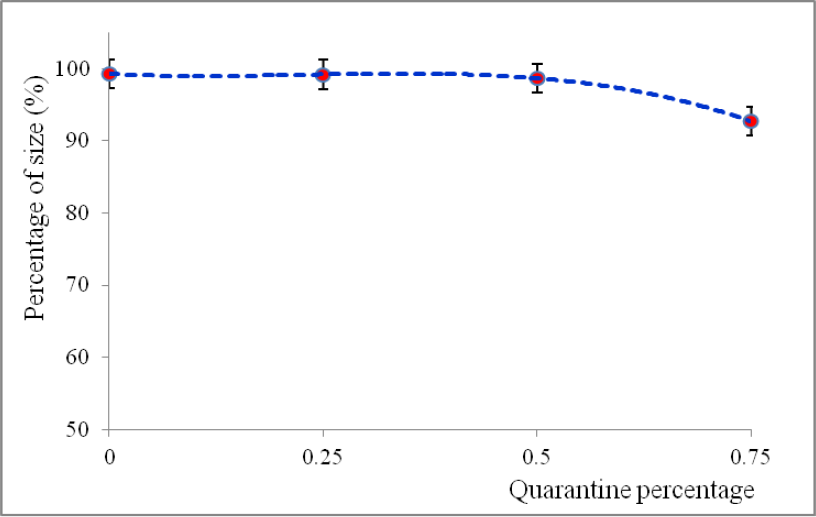
The size percentage as a function of quarantine degree. The dotted line represents a 2^nd^ degree polynomial fit.

**Figure 6:**
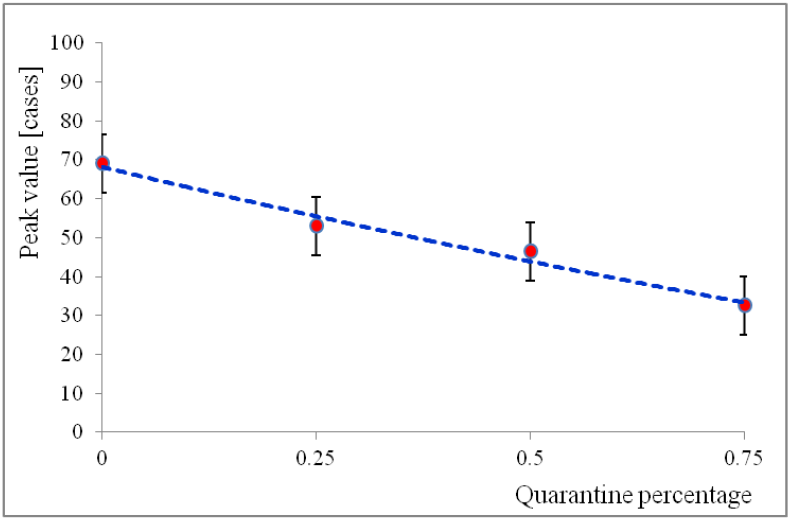
The peak value of DNC as a function of quarantine degree.The dotted line represents a 2^nd^ degree polynomial fit.

**Figure 7:**
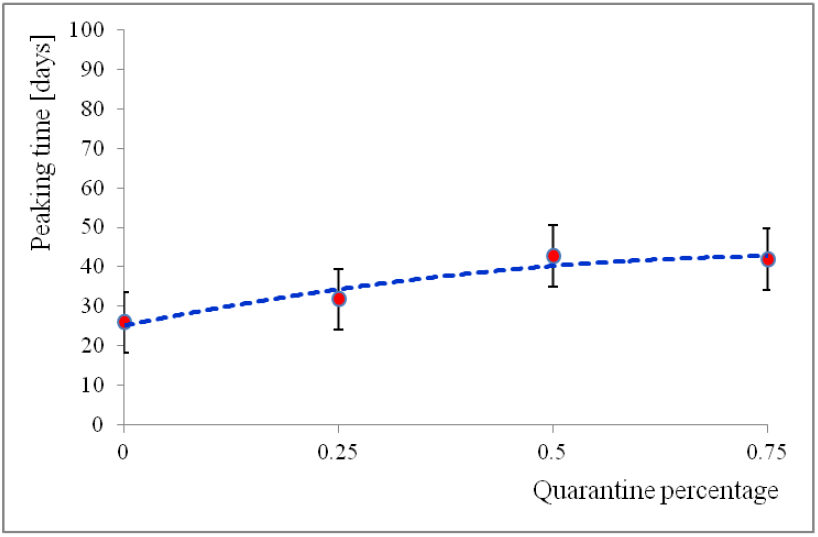
The peaking time of DNC as a function of quarantine degree. The dotted line represents a 2^nd^ degree polynomial fit.

**Figure 8:**
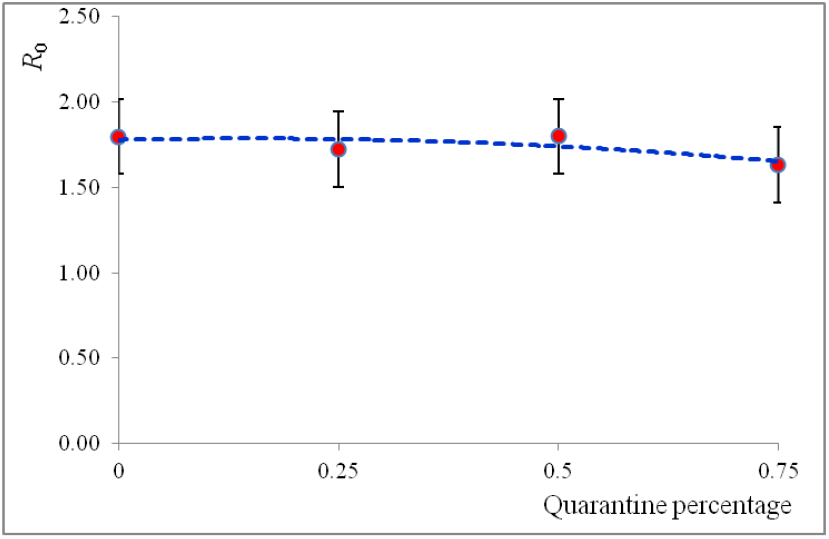
The basic reproductive number as a function of quarantine degree. The dotted line represents a 2^nd^ degree polynomial fit.

Observing these results we find some interesting consequences of the quarantine as follows: The percentage of size shows a very small decrease of 6% within the time scale under study. Also, the greater the degree of quarantine the greater the decrease in the DNC peak value, while the peaking time extended accordingly. Concerning the basic reproductive number, *R*_0_, we observe that it shows negligible variation compared with the statistical errors and thus we can consider it to be unchanged. As we know from the SIR-based epidemic models, *R*_0_ is a fundamental quantity related to the two basic parameters, the transmission rate and the recovering rate and therefore can not be affected seriously by the quarantine alone. The overall variations, from 0 to 75%, due to the quarantine are: the DNC peak value is decreased by 53%, while the DNC peaking time is extended by 61%.

### 4.3 Results analysing real epidemiological data

To compare the results of the MC simulation with real epidemiological data we have chosen a country where the raw data and any parametrization of them present a “picture” very close to the theoretical as well as having relatively small fluctuations. This country is Switzerland where, besides the above characteristics, the compliance with the measures is largely secured. Therefore, it can be considered to be a well reference for comparison. In Fig. 9 and Fig. 10 the NDC data parametrized by the LPE-SG model and the *R*_e_ are shown respectively [27]. The results of the MC simulation we developed show a generic consistency with the real ones, by means of the DNC shape, peaking time, overall time scale of the epidemic and the *R*_e_ drop-off shape.

**Figure 9:**
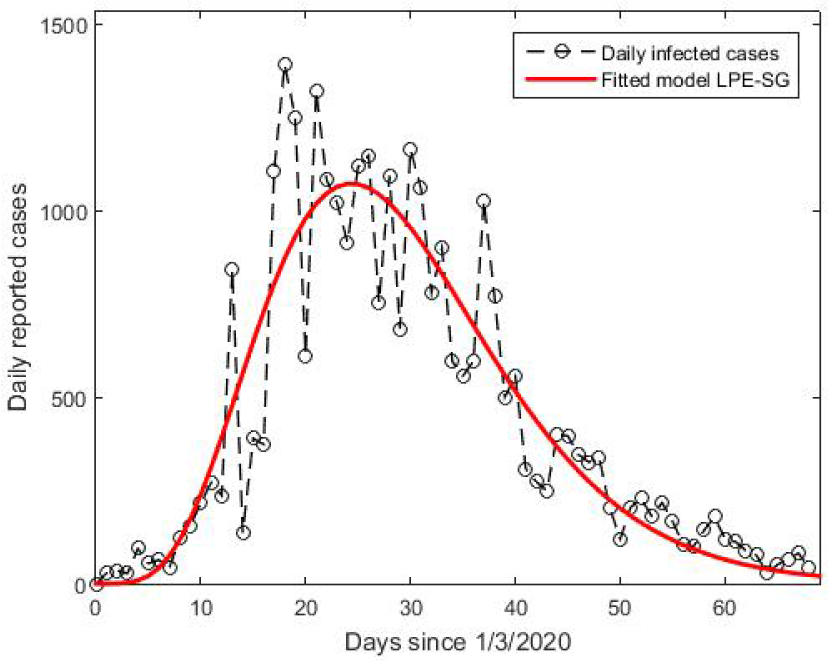
The epidemiological data (DNC) for the first phase of epidemic outbreak in Switzerland since 1/3/2020, obtained from [5].

**Figure 10:**
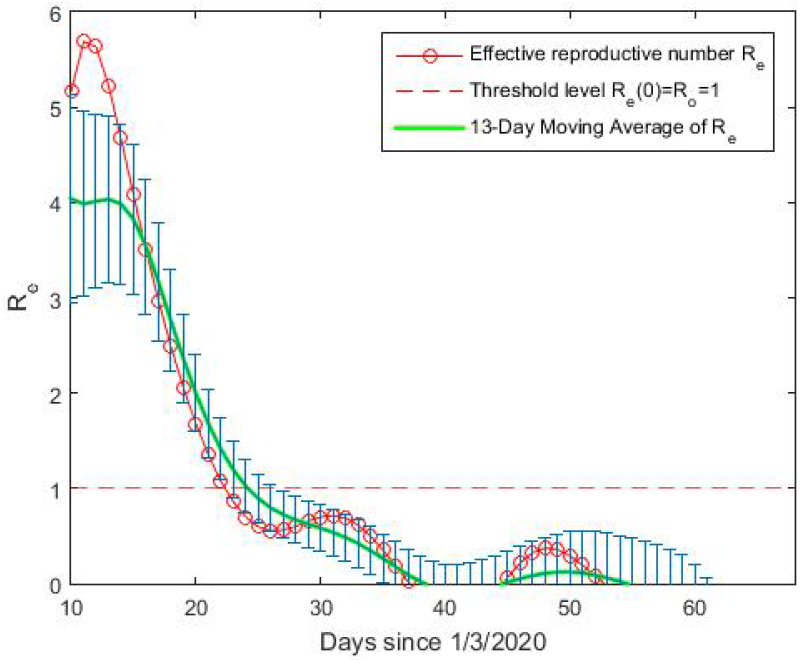
The effective reproductive number as a function of time for the first phase of epidemic outbreak in Switzerland, reproduced as given in [21].

## Conclusions

A Monte Carlo simulation has been developed to study the spread dynamics of the new COVID-19 disease. The novel approach we followed in methodology was based on fundamental mechanisms during the spread of the epidemic. One issue of key importance was the realistic selection of statistical distributions so that the results of the simulation correspond to those previously recorded to date in various countries during the first and second “wave” of the epidemic. This task was faced by studying the appropriate statistical distributions and their associated parameters. Moreover, we described the mathematical process for determining the effective reproductive number in good approximation, either by the simulated or by the real epidemiological data. This quantity is very important in playing the role of “figure of merit” helping to survey and understand the dynamics of the epidemic spread. We also studied the effect of quarantine at different levels of its implementation on the shape of the epidemic curve and its parameters. In addition, we present some indicative real epidemic results analysed using a specific parametrization model.

## Data Availability

The data concern the parameters and applied on epidemiological data.

## Acknowledgements

We gratefully thank Mrs. Barbara Theodoropoulou, experienced English teacher, for her willing support in the language proofreading.

